# High prevalence of SARS-CoV-2 swab positivity and increasing R number in England during October 2020: REACT-1 round 6 interim report

**DOI:** 10.1101/2020.10.30.20223123

**Authors:** Steven Riley, Kylie E. C. Ainslie, Oliver Eales, Caroline E. Walters, Haowei Wang, Christina Atchison, Claudio Fronterre, Peter J. Diggle, Deborah Ashby, Christl A. Donnelly, Graham Cooke, Wendy Barclay, Helen Ward, Ara Darzi, Paul Elliott

## Abstract

**Background:** REACT-1 measures prevalence of SARS-CoV-2 infection in representative samples of the population in England using PCR testing from self-administered nose and throat swabs. Here we report interim results for round 6 of observations for swabs collected from the 16th to 25th October 2020 inclusive.

**Methods:** REACT-1 round 6 aims to collect data and swab results from 160,000 people aged 5 and above. Here we report results from the first 86,000 individuals. We estimate prevalence of PCR-confirmed SARS-CoV-2 infection, reproduction numbers (R) and temporal trends using exponential growth or decay models. Prevalence estimates are presented both unweighted and weighted to be representative of the population of England, accounting for response rate, region, deprivation and ethnicity. We compare these interim results with data from round 5, based on swabs collected from 18th September to 5th October 2020 inclusive.

**Results:** Overall prevalence of infection in the community in England was 1.28% or 128 people per 10,000, up from 60 per 10,000 in the previous round. Infections were doubling every 9.0 (6.1, 18) days with a national reproduction number (R) estimated at 1.56 (1.27, 1.88) compared to 1.16 (1.05, 1.27) in the previous round. Prevalence of infection was highest in Yorkshire and The Humber at 2.72% (2.12%, 3.50%), up from 0.84% (0.60%, 1.17%), and the North West at 2.27% (1.90%, 2.72%), up from 1.21% (1.01%, 1.46%), and lowest in South East at 0.55% (0.45%, 0.68%), up from 0.29% (0.23%, 0.37%). Clustering of cases was more prevalent in Lancashire, Manchester, Liverpool and West Yorkshire, West Midlands and East Midlands. Interim estimates of R were above 2 in the South East, East of England, London and South West, but with wide confidence intervals. Nationally, prevalence increased across all age groups with the greatest increase in those aged 55-64 at 1.20% (0.99%, 1.46%), up 3-fold from 0.37% (0.30%, 0.46%). In those aged over 65, prevalence was 0.81% (0.58%, 0.96%) up 2-fold from 0.35% (0.28%, 0.43%). Prevalence remained highest in 18 to 24-year olds at 2.25% (1.47%, 3.42%).

**Conclusion:** The co-occurrence of high prevalence and rapid growth means that the second wave of the epidemic in England has now reached a critical stage. Whether via regional or national measures, it is now time-critical to control the virus and turn R below one if further hospital admissions and deaths from COVID-19 are to be avoided.

## Introduction

The COVID-19 pandemic in England has followed two distinct patterns: a first wave in March-April 2020 where a large peak of infections was followed by a sharp fall during lockdown, with a fall in infections continuing until early August [1]. Since then there has been a continuous rise in infections (‘second wave’), with reproduction number (R) persistently above one, leading to high prevalence of infections, particularly in northern regions [2]. The REal-time Assessment of Community Transmission-1 (REACT-1) study was established to track the course of the epidemic in England since May 2020 [3,4]. We recently reported results of round 5 of the study which obtained swabs for rt-PCR from 18th September to 5th October 2020 [2]. That study reported a national prevalence of 0.60% with particularly high prevalence and growth in the north of the country. Here we report interim results for round 6, for swabs obtained from 16th to 25th October.

## Methods

REACT-1 methods have been described previously [3,4]. In brief, we obtain a nose and throat swab from non-overlapping random samples of the population of England (stratified by the 315 lower-tier local authorities) at ages five years and above, using the National Health Service (NHS) list of GP registered patients as the sampling frame. Participants also complete a questionnaire. Swabs are picked up by courier from the participant’s home and kept cold until PCR testing to maintain sample integrity. We obtain estimates of prevalence (and 95% confidence intervals) both unweighted and weighted to be representative of the population of England, accounting for response rate, region, deprivation and ethnicity. We fit exponential growth and decay models to analyse time trends in swab positivity both across subsequent rounds of the study and latterly (where numbers of positive swabs have been higher) within each round. We also performed multivariable logistic regression to determine the relationship between different covariates on swab positivity.

We obtained research ethics approval from the South Central-Berkshire B Research Ethics Committee (IRAS ID: 283787).

## Results

We found 863 positives from 85,971 swabs giving an unweighted prevalence of 1.00% (95% CI, 0.94%, 1.07%) and a weighted prevalence of 1.28% (1.15%, 1.41%). The weighted prevalence estimate is more than double that of 0.60% (0.55%, 0.71%) obtained in round 5 (Table 1).

**Table 1.** Unweighted and weighted prevalence of swab-positivity across five complete rounds of REACT-1 and partial data from round 6.

| Round | Tested swabs | Positive swabs | Unweighted prevalence (95% CI) | Weighted prevalence (95% CI) | First sample | Last sample |
| --- | --- | --- | --- | --- | --- | --- |
| 1 | 120,620 | 159 | 0.13% (0.11%, 0.15%) | 0.16% (0.13%, 0.19%) | 1/5/2020 | 1/6/2020 |
| 2 | 159,199 | 123 | 0.077% (0.065%, 0.092%) | 0.088% (0.068%, 0.11%) | 19/6/2020 | 7/7/2020 |
| 3 | 162,821 | 54 | 0.033% (0.025%, 0.043%) | 0.040% (0.027%, 0.053%) | 24/7/2020 | 11/8/2020 |
| 4 | 154,325 | 137 | 0.089% (0.075%, 0.11%) | 0.13% (0.096%, 0.15%) | 20/8/2020 | 8/9/2020 |
| 5 | 174,949 | 824 | 0.47% (0.44%, 0.50%) | 0.60% (0.55%, 0.71%) | 18/9/2020 | 5/10/2020 |
| 6* | 85,971 | 863 | 1.00% (0.94%, 1.07%) | 1.28% (1.15%, 1.41%) | 16/10/2020 | 25/10/2020 |

\*Not yet complete

The increase in prevalence gave an average R number over the period of rounds 5 and 6 of 1.20 (1.18,1.23) (Table 2, Figure 1A, Figure 2). This R number from rounds 5 to 6 is similar to that reported for round 5 alone of 1.16 (1.05, 1.27).

**Table 2.** National estimates of growth rate, doubling time and reproduction number for rounds 5 and 6 together and for round 6 alone.

| Data used | Outcome | Growth rate $r$ (1/days) | R rate | $p(R>1)$ | Doubling (+) / halving (-) time (days) |
| --- | --- | --- | --- | --- | --- |
| Rounds 5 and 6 | All positives | 0.031 ( 0.027 , 0.034 ) | 1.20 ( 1.18 , 1.23 ) | 1.000 | 22.7 ( 26.0 , 20.1 ) |
|  | Non-symptomatics | 0.034 ( 0.028 , 0.040 ) | 1.23 ( 1.19 , 1.27 ) | 1.000 | 20.5 ( 24.9 , 17.4 ) |
|  | Positive for both E and N genes | 0.024 ( 0.019 , 0.029 ) | 1.16 ( 1.13 , 1.19 ) | 1.000 | 29.2 ( 36.2 , 24.3 ) |
|  | Positive for both E and N genes or positive only for N gene with CT 35 or less | 0.029 ( 0.025 , 0.033 ) | 1.19 ( 1.16 , 1.22 ) | 1.000 | 23.9 ( 27.9 , 20.9 ) |
| Round 6 | All positives | 0.077 ( 0.039 , 0.114 ) | 1.56 ( 1.27 , 1.88 ) | 1.000 | 9.0 ( 17.7 , 6.1 ) |
|  | Non-symptomatics | 0.139 ( 0.083 , 0.194 ) | 2.12 ( 1.61 , 2.68 ) | 1.000 | 5.0 ( 8.4 , 3.6 ) |
|  | Positive for both E and N genes | 0.055 ( 0.009 , 0.102 ) | 1.39 ( 1.06 , 1.77 ) | 0.991 | 12.5 ( 77.8 , 6.8 ) |
|  | Positive for both E and N genes or positive only for N gene with CT 35 or less | 0.057 ( 0.017 , 0.098 ) | 1.40 ( 1.11 , 1.73 ) | 0.998 | 12.2 ( 40.2 , 7.1 ) |

**Figure 1.**
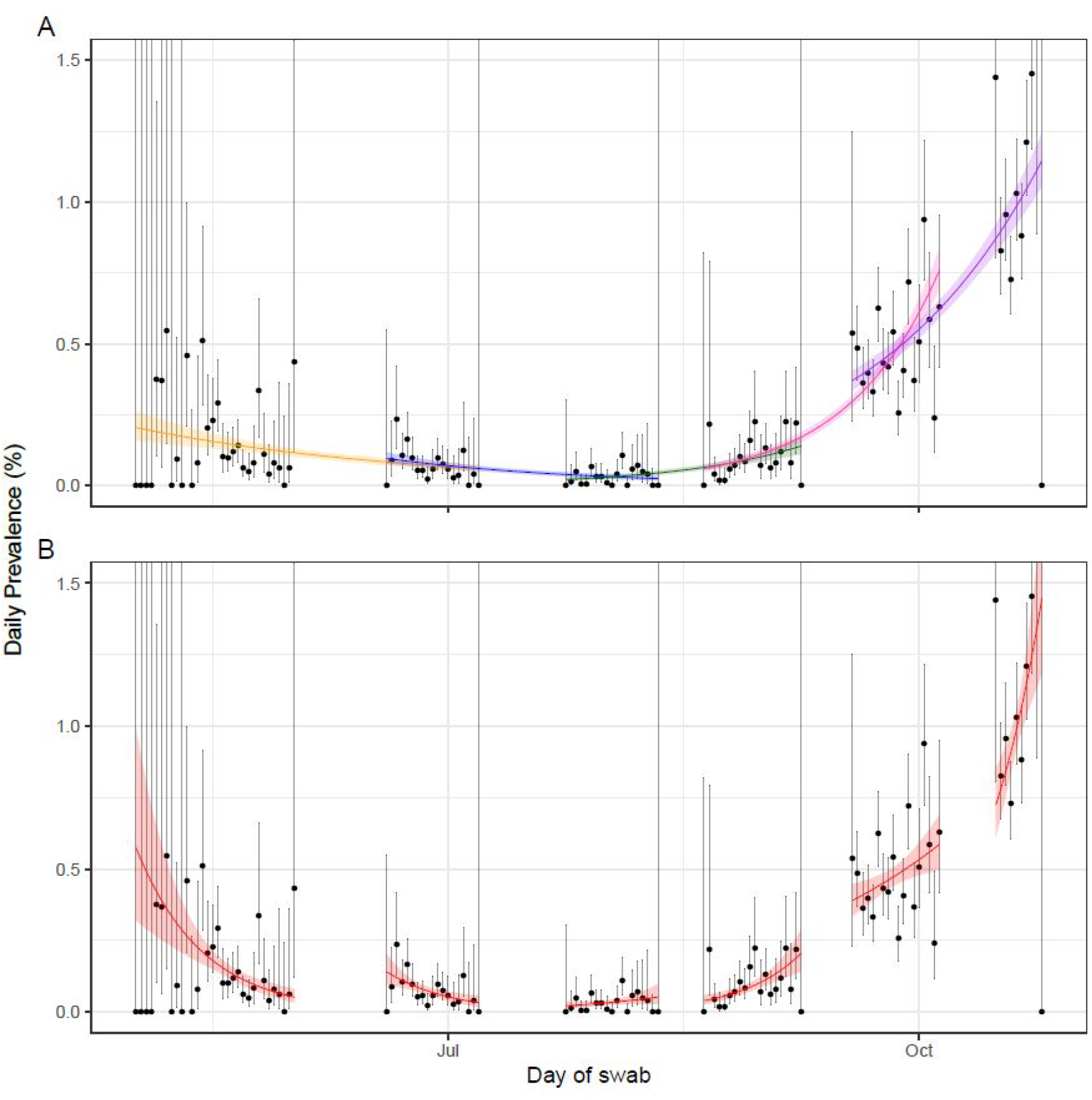
Constant growth rate models fit to REACT-1 data for sequential and individual rounds. **A** models fit to REACT-1 data for sequential rounds; 1 and 2 (yellow), 2 and 3 (blue), 3 and 4 (green), 4 and 5 (pink), and 5 and 6 (purple). **B** models fit to individual rounds only (red). Vertical lines show 95% confidence intervals for observed prevalence (black points). Shaded regions show 95% posterior credible intervals for growth models. Note that of the 867,700 swab tests only 849,544 had a date available (2199 of 2294 positives) and so could be included in this temporal analysis.

**Figure 2.**
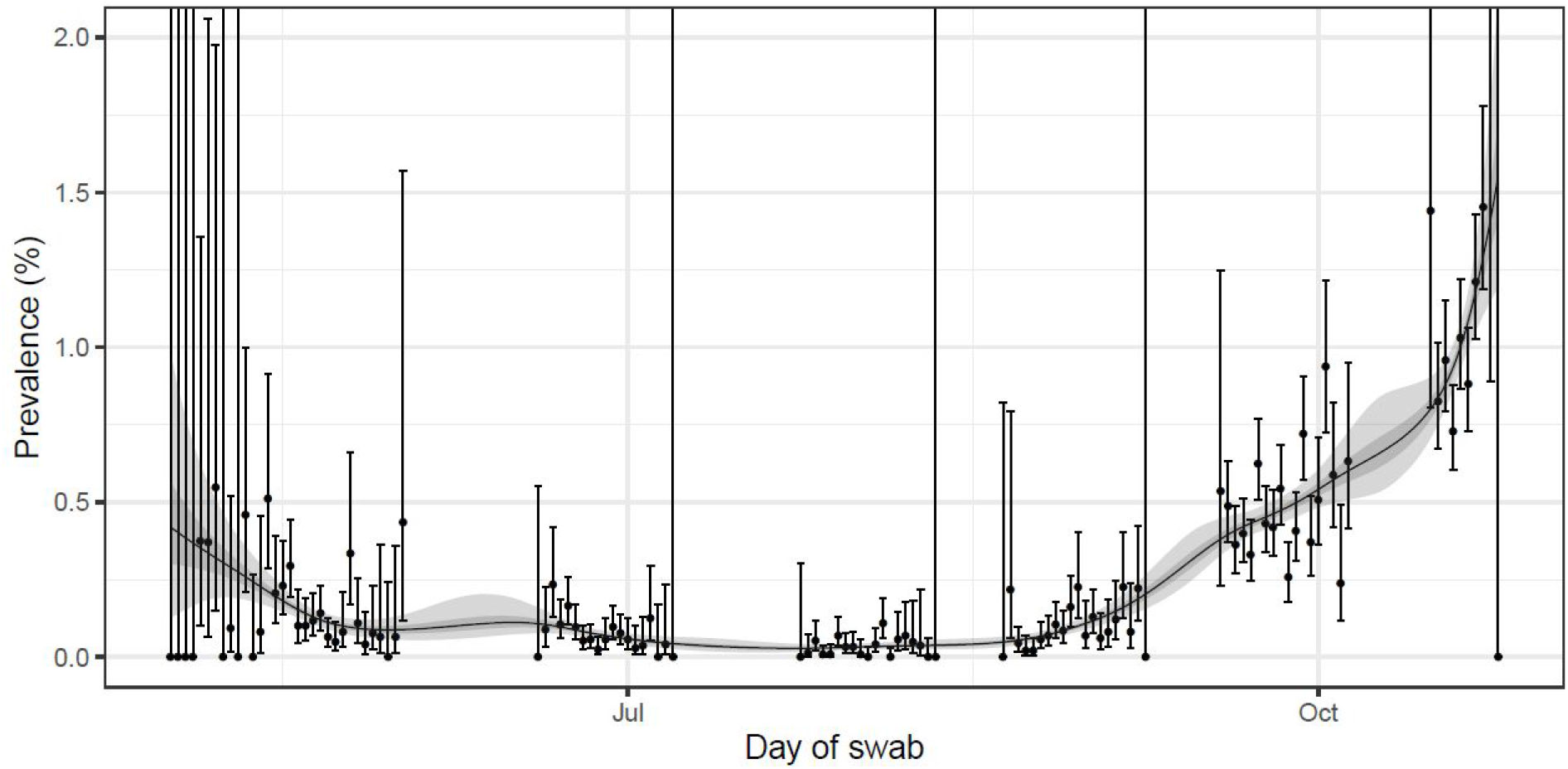
Prevalence of swab-positivity estimated for the full period of the study with central 50% and 95% posterior credible intervals. 849,544 of the total 867,700 swab tests had a date available (2199 of 2294 positives) and so could be included in this temporal analysis. We fit a Bayesian P-spline model [7] to the daily data using a No U-Turns Sampler in logit space. The data is split into 36 regularly sized segments, controlled by 37 regularly placed knots (chosen so that each segment is approximately 5 days), with further knots defined beyond the boundaries of the period in order to remove edge effects. A system of 4th order B-splines are defined over the knots and the model consists of a linear combination of these B-splines. Overfitting is prevented through the inclusion of a second-order random-walk prior on the coefficients of the B-splines. This has the effect of penalising any changes in the growth rate; the magnitude of this effect is controlled by a further parameter.

However, data from round 6 alone suggest a more recent increase in the national R. Within round 6 we estimate a national doubling time of 9.0 (6.1, 17.7) days corresponding to an R of 1.56 (1.27, 1.88) (Table 2, Figure 1B). Although based on a short period of time, the 95% credible interval for this estimate of national R for round 6 alone does not overlap with our estimate for rounds 5 and 6 together. Also, our estimate for round 6 alone is substantially higher than that estimated for round 5 alone. Sensitivity analyses showed similar R estimates for rounds 5 and 6 together and for round 6 alone for double gene target positives, for positive with a lower CT value cut-off for the N gene, and for non-symptomatic participants (Table 2).

There has been an increase in prevalence in all regions since round 5 (Table 3a, Table 3b, Figure 3). Weighted prevalence was highest in the North: Yorkshire and The Humber at 2.72% (2.12%, 3.50%) and North West at 2.27% (1.90%, 2.72%). Weighted prevalence was lowest in the South East at 0.55% (0.45%, 0.68%).

**Table 3a.** Unweighted prevalence of swab-positivity by variable and category for rounds 5 and 6 (partial) of REACT-1.

| Variable | Category | Round 5 |  |  | Round 6 |  |  |
| --- | --- | --- | --- | --- | --- | --- | --- |
|  |  | Positive | Total | Prevalence | Positive | Total | Prevalence |
| Gender | Male | 387 | 78,565 | 0.49% ( 0.45% , 0.54% ) | 387 | 39,319 | 0.98% ( 0.89% , 1.09% ) |
|  | Female | 437 | 96,380 | 0.45% ( 0.41% , 0.50% ) | 476 | 46,648 | 1.02% ( 0.93% , 1.12% ) |
| Age | 05-12 | 59 | 13,771 | 0.43% ( 0.33% , 0.55% ) | 55 | 5,513 | 1.00% ( 0.77% , 1.30% ) |
|  | 13-17 | 59 | 10,480 | 0.56% ( 0.44% , 0.73% ) | 60 | 5,169 | 1.16% ( 0.90% , 1.49% ) |
|  | 18-24 | 83 | 6,852 | 1.21% ( 0.98% , 1.50% ) | 53 | 3,091 | 1.71% ( 1.31% , 2.24% ) |
|  | 25-34 | 86 | 15,683 | 0.55% ( 0.44% , 0.68% ) | 97 | 7,186 | 1.35% ( 1.11% , 1.64% ) |
|  | 35-44 | 121 | 22,954 | 0.53% ( 0.44% , 0.63% ) | 98 | 10,079 | 0.97% ( 0.80% , 1.18% ) |
|  | 45-54 | 157 | 29,380 | 0.53% ( 0.46% , 0.62% ) | 137 | 13,009 | 1.05% ( 0.89% , 1.24% ) |
|  | 55-64 | 116 | 31,885 | 0.36% ( 0.30% , 0.44% ) | 167 | 15,768 | 1.06% ( 0.91% , 1.23% ) |
|  | 65+ | 143 | 43,944 | 0.33% ( 0.28% , 0.38% ) | 196 | 26,156 | 0.75% ( 0.65% , 0.86% ) |
| Region | South East | 104 | 39,496 | 0.26% ( 0.22% , 0.32% ) | 106 | 19,367 | 0.55% ( 0.45% , 0.66% ) |
|  | North East | 61 | 6,702 | 0.91% ( 0.71% , 1.17% ) | 45 | 3,323 | 1.35% ( 1.01% , 1.81% ) |
|  | North West | 199 | 19,468 | 1.02% ( 0.89% , 1.17% ) | 192 | 10,035 | 1.91% ( 1.66% , 2.20% ) |
|  | Yorkshire and The Humber | 74 | 11,658 | 0.63% ( 0.51% , 0.80% ) | 113 | 6,179 | 1.83% ( 1.52% , 2.19% ) |
|  | East Midlands | 101 | 21,897 | 0.46% ( 0.38% , 0.56% ) | 128 | 11,461 | 1.12% ( 0.94% , 1.33% ) |
|  | West Midlands | 70 | 16,343 | 0.43% ( 0.34% , 0.54% ) | 90 | 8,005 | 1.12% ( 0.92% , 1.38% ) |
|  | East of England | 81 | 25,625 | 0.32% ( 0.25% , 0.39% ) | 74 | 12,612 | 0.59% ( 0.47% , 0.74% ) |
|  | London | 79 | 17,500 | 0.45% ( 0.36% , 0.56% ) | 61 | 7,299 | 0.84% ( 0.65% , 1.07% ) |
|  | South West | 55 | 16,260 | 0.34% ( 0.26% , 0.44% ) | 54 | 7,690 | 0.70% ( 0.54% , 0.92% ) |
| Employment type | Health care or care home worker | 51 | 9,620 | 0.53% ( 0.40% , 0.70% ) | 54 | 4,069 | 1.33% ( 1.02% , 1.73% ) |
|  | Other essential/key worker | 154 | 28,846 | 0.53% ( 0.46% , 0.62% ) | 169 | 12,503 | 1.35% ( 1.16% , 1.57% ) |
|  | Other worker | 338 | 72,175 | 0.47% ( 0.42% , 0.52% ) | 366 | 34,262 | 1.07% ( 0.96% , 1.18% ) |
|  | Not full-time, part-time, or self-employed | 256 | 59,762 | 0.43% ( 0.38% , 0.48% ) | 256 | 33,136 | 0.77% ( 0.68% , 0.87% ) |
| Ethnic group | White | 707 | 157,645 | 0.45% ( 0.42% , 0.48% ) | 793 | 79,406 | 1.00% ( 0.93% , 1.07% ) |
|  | Asian | 71 | 7,907 | 0.90% ( 0.71% , 1.13% ) | 35 | 2,863 | 1.22% ( 0.88% , 1.70% ) |
|  | Black | 16 | 2,194 | 0.73% ( 0.45% , 1.18% ) | 7 | 603 | 1.16% ( 0.56% , 2.38% ) |
|  | Mixed | 11 | 3,080 | 0.36% ( 0.20% , 0.64% ) | 14 | 1,326 | 1.06% ( 0.63% , 1.76% ) |
|  | Other | 7 | 1,439 | 0.49% ( 0.24% , 1.00% ) | 6 | 536 | 1.12% ( 0.51% , 2.42% ) |
| Household size | 1 | 88 | 24,916 | 0.35% ( 0.29% , 0.43% ) | 106 | 12,368 | 0.86% ( 0.71% , 1.04% ) |
|  | 2 | 227 | 61,717 | 0.37% ( 0.32% , 0.42% ) | 306 | 34,116 | 0.90% ( 0.80% , 1.00% ) |
|  | 3 | 172 | 32,395 | 0.53% ( 0.46% , 0.62% ) | 180 | 15,409 | 1.17% ( 1.01% , 1.35% ) |
|  | 4 | 211 | 37,231 | 0.57% ( 0.50% , 0.65% ) | 161 | 16,329 | 0.99% ( 0.85% , 1.15% ) |
|  | 5 | 77 | 13,145 | 0.59% ( 0.47% , 0.73% ) | 76 | 5,438 | 1.40% ( 1.12% , 1.75% ) |
|  | 6 | 31 | 3,845 | 0.81% ( 0.57% , 1.14% ) | 27 | 1,632 | 1.65% ( 1.14% , 2.40% ) |
|  | 7+ | 18 | 1,700 | 1.06% ( 0.67% , 1.67% ) | 7 | 679 | 1.03% ( 0.50% , 2.11% ) |
| COVID case contact | No | 556 | 139,853 | 0.40% ( 0.37% , 0.43% ) | 529 | 67,864 | 0.78% ( 0.72% , 0.85% ) |
|  | Yes, contact with a confirmed/tested COVID-19 case | 102 | 1,566 | 6.51% ( 5.39% , 7.85% ) | 147 | 1,787 | 8.23% ( 7.04% , 9.59% ) |
|  | Yes, contact with a suspected COVID-19 case | 17 | 1,487 | 1.14% ( 0.72% , 1.82% ) | 22 | 842 | 2.61% ( 1.73% , 3.92% ) |
| Deprivation | 1 Most deprived | 135 | 17,641 | 0.77% ( 0.65% , 0.91% ) | 119 | 7,919 | 1.50% ( 1.26% , 1.80% ) |
|  | 2 | 129 | 27,483 | 0.47% ( 0.40% , 0.56% ) | 143 | 12,750 | 1.12% ( 0.95% , 1.32% ) |
|  | 3 | 146 | 37,427 | 0.39% ( 0.33% , 0.46% ) | 176 | 18,292 | 0.96% ( 0.83% , 1.11% ) |
|  | 4 | 190 | 43,109 | 0.44% ( 0.38% , 0.51% ) | 199 | 21,749 | 0.92% ( 0.80% , 1.05% ) |
|  | 5 Least deprived | 224 | 49,289 | 0.45% ( 0.40% , 0.52% ) | 226 | 25,261 | 0.89% ( 0.79% , 1.02% ) |

**Table 3b.**
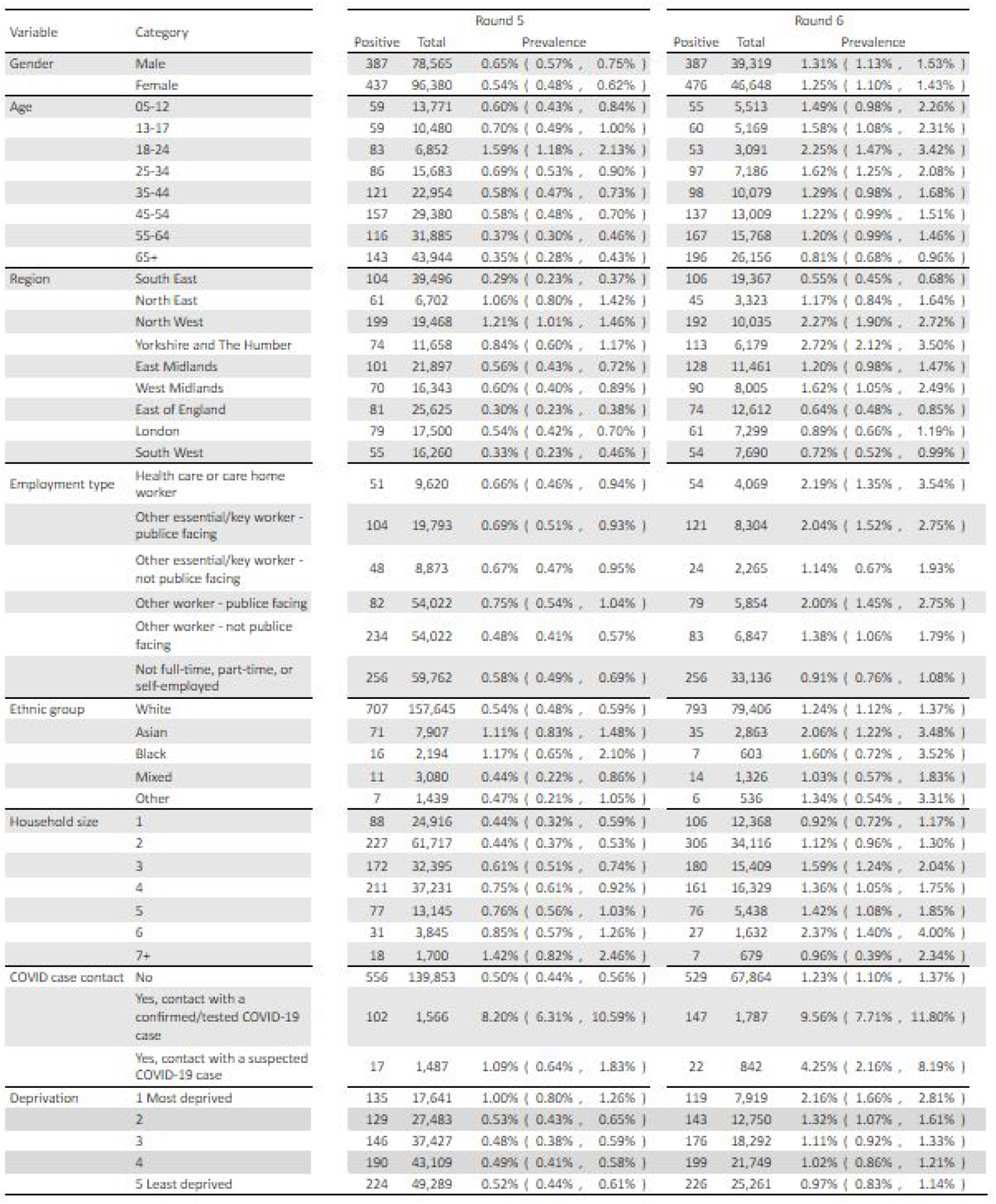
Weighted prevalence of swab-positivity by variable and category for rounds 5 and 6 (partial) of REACT-1.

**Figure 3.**
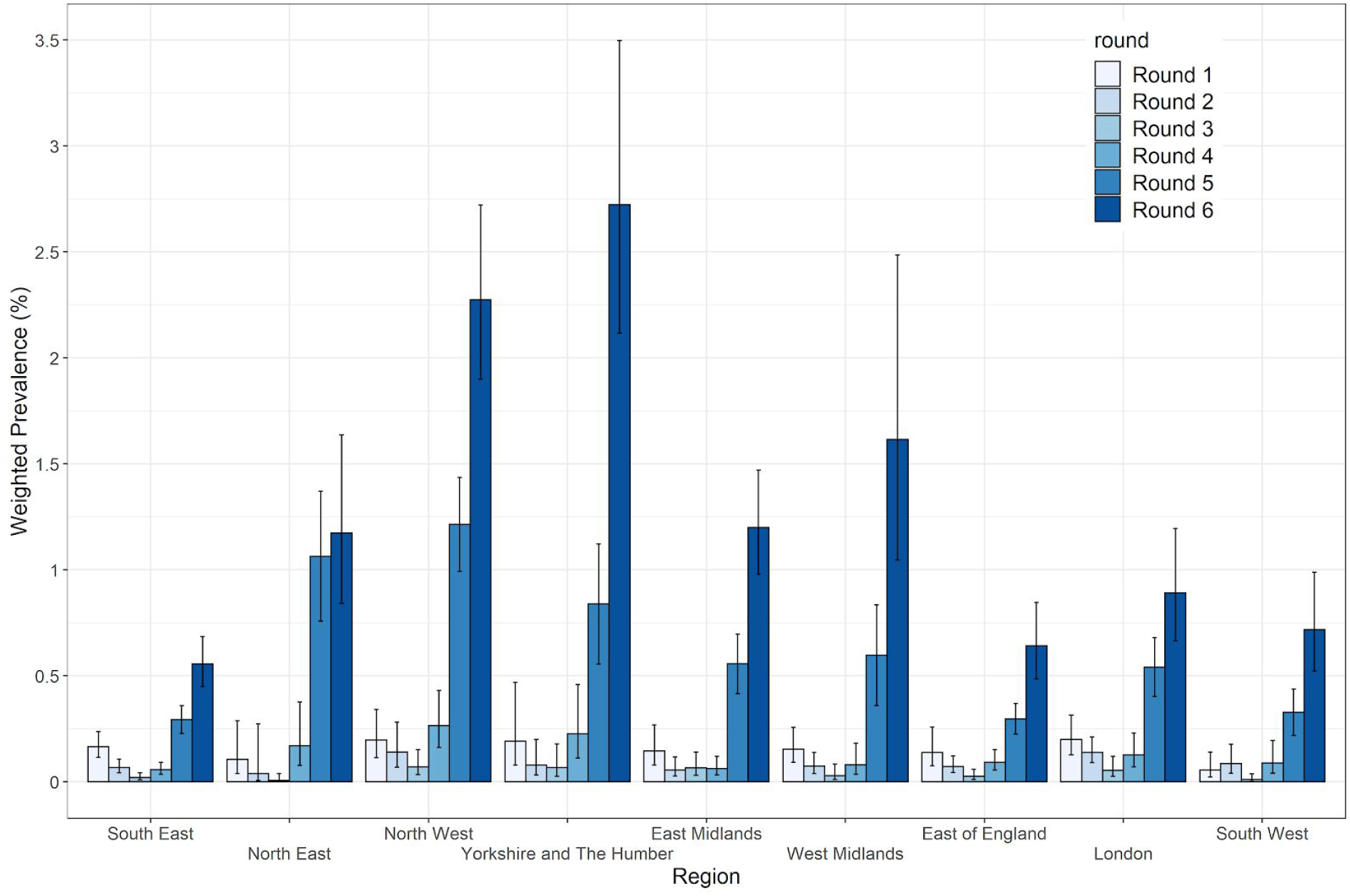
Weighted prevalence and 95% confidence intervals of swab positivity by region by round.

However, based on estimates of regional R for round 6 alone, epidemic growth is no longer fastest in the North (Table 4, Figure 4). Interim point estimates of R were above 2 in the South East, East of England, London and South West, but with wide confidence intervals (Table 4). Further, we found a 99% probability that R values were greater than 1 in London, South East, and East of England. We found between 95% and 99% probability that R values were greater than 1 in the South West and West Midlands.

**Table 4.** Regional estimates of growth rate, doubling time and reproduction number for rounds 5 and 6 together and for round 6 alone.

| Data used | Region | Growth rate $r$ (1/days) | R number | $p(R>1)$ | Doubling (+) / halving (-) time (days) |
| --- | --- | --- | --- | --- | --- |
| Rounds 5 and 6 | South East | 0.032 ( 0.021 , 0.043 ) | 1.21 ( 1.14 , 1.30 ) | 1.000 | 21.7 ( 33.1 , 16.1 ) |
|  | North East | 0.011 ( -0.005 , 0.026 ) | 1.07 ( 0.97 , 1.17 ) | 0.911 | 63.6 ( -149.2 , 26.5 ) |
|  | North West | 0.027 ( 0.019 , 0.035 ) | 1.18 ( 1.12 , 1.24 ) | 1.000 | 25.9 ( 37.2 , 19.7 ) |
|  | Yorkshire and The Humber | 0.043 ( 0.031 , 0.055 ) | 1.29 ( 1.21 , 1.39 ) | 1.000 | 16.1 ( 22.7 , 12.5 ) |
|  | East Midlands | 0.036 ( 0.026 , 0.047 ) | 1.25 ( 1.17 , 1.32 ) | 1.000 | 19.1 ( 26.8 , 14.8 ) |
|  | West Midlands | 0.042 ( 0.029 , 0.056 ) | 1.29 ( 1.20 , 1.39 ) | 1.000 | 16.3 ( 23.6 , 12.5 ) |
|  | East of England | 0.025 ( 0.013 , 0.038 ) | 1.17 ( 1.08 , 1.26 ) | 1.000 | 27.4 ( 55.1 , 18.2 ) |
|  | London | 0.025 ( 0.011 , 0.039 ) | 1.17 ( 1.07 , 1.26 ) | 1.000 | 27.9 ( 61.8 , 17.8 ) |
|  | South West | 0.029 ( 0.013 , 0.045 ) | 1.20 ( 1.09 , 1.31 ) | 1.000 | 23.7 ( 52.2 , 15.4 ) |
| Round 6 | South East | 0.162 ( 0.052 , 0.271 ) | 2.34 ( 1.36 , 3.61 ) | 0.999 | 4.3 ( 13.4 , 2.6 ) |
|  | North East | -0.079 ( -0.248 , 0.085 ) | 0.57 ( 0.07 , 1.63 ) | 0.170 | -8.8 ( -2.8 , 8.1 ) |
|  | North West | 0.031 ( -0.049 , 0.110 ) | 1.21 ( 0.72 , 1.84 ) | 0.774 | 22.6 ( -14.3 , 6.3 ) |
|  | Yorkshire and The Humber | 0.075 ( -0.026 , 0.174 ) | 1.54 ( 0.84 , 2.47 ) | 0.927 | 9.2 ( -27.0 , 4.0 ) |
|  | East Midlands | 0.057 ( -0.033 , 0.151 ) | 1.40 ( 0.80 , 2.23 ) | 0.894 | 12.1 ( -20.9 , 4.6 ) |
|  | West Midlands | 0.107 ( -0.007 , 0.220 ) | 1.81 ( 0.96 , 2.98 ) | 0.968 | 6.5 ( -104.6 , 3.1 ) |
|  | East of England | 0.146 ( 0.023 , 0.278 ) | 2.18 ( 1.15 , 3.70 ) | 0.990 | 4.7 ( 30.0 , 2.5 ) |
|  | London | 0.210 ( 0.066 , 0.359 ) | 2.86 ( 1.47 , 4.87 ) | 0.998 | 3.3 ( 10.5 , 1.9 ) |
|  | South West | 0.133 ( -0.018 , 0.284 ) | 2.06 ( 0.89 , 3.79 ) | 0.957 | 5.2 ( -38.3 , 2.4 ) |

**Figure 4.**
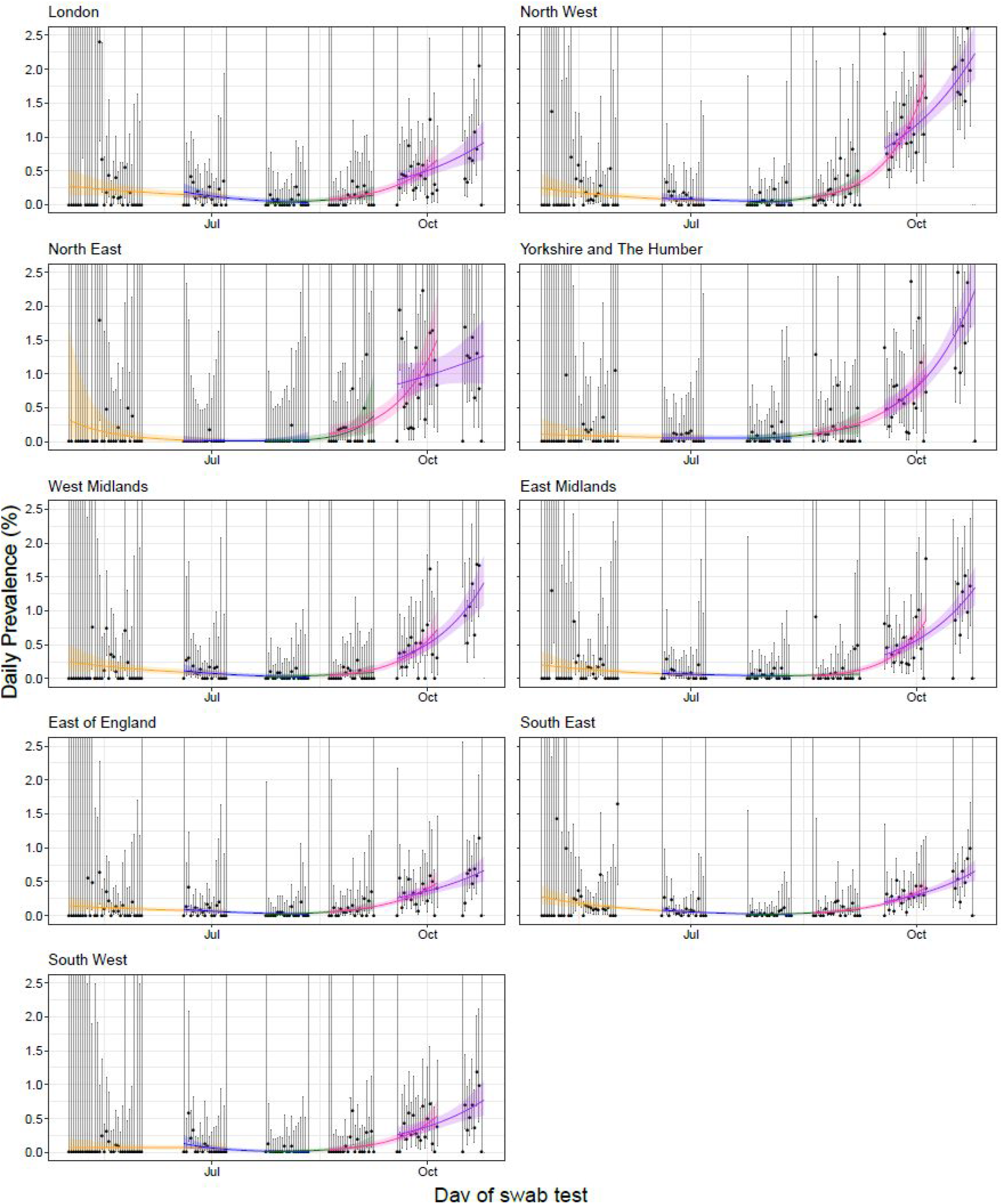
Constant growth rate models fit to regions for REACT-1 data for sequential rounds: 1 and 2 (yellow), 2 and 3 (blue), 3 and 4 (green), 4 and 5 (pink), and 5 and 6 (purple). Vertical lines show 95% confidence intervals for observed prevalence (black points). Shaded regions show 95% posterior credible intervals for growth models. Note that of the 867,700 swab tests only 849,544 had a date available (2199 of 2294 positives) and so could be included in this temporal analysis.

Nationally, prevalence increased across all age groups with the greatest increase in those aged 55-64 at 1.20% (0.99%, 1.46%), up 3-fold from 0.37% (0.30%, 0.46%) (Table 3b, Figure 5). In those aged over 65, prevalence was 0.81% (0.58%, 0.96%) up 2-fold from 0.35% (0.28%, 0.43%). The highest weighted prevalence was seen in the 18 to 24 year olds at 2.25% (1.47%, 3.42), which has increased from 1.59% (1.18%, 2.13%) for round 5.

**Figure 5.**
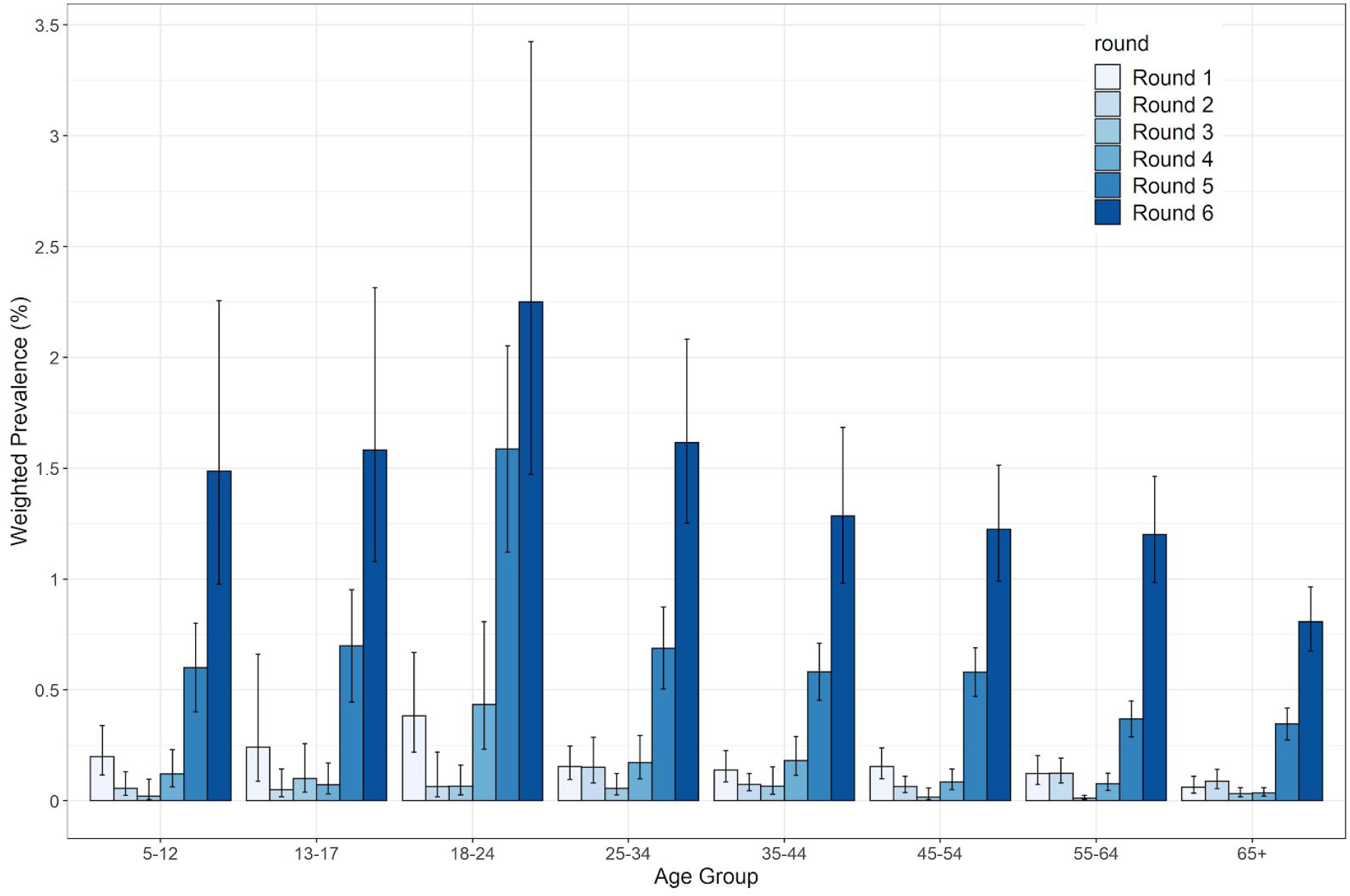
Weighted prevalence and 95% confidence intervals of swab-positivity by age group by round.

**Figure 6.**
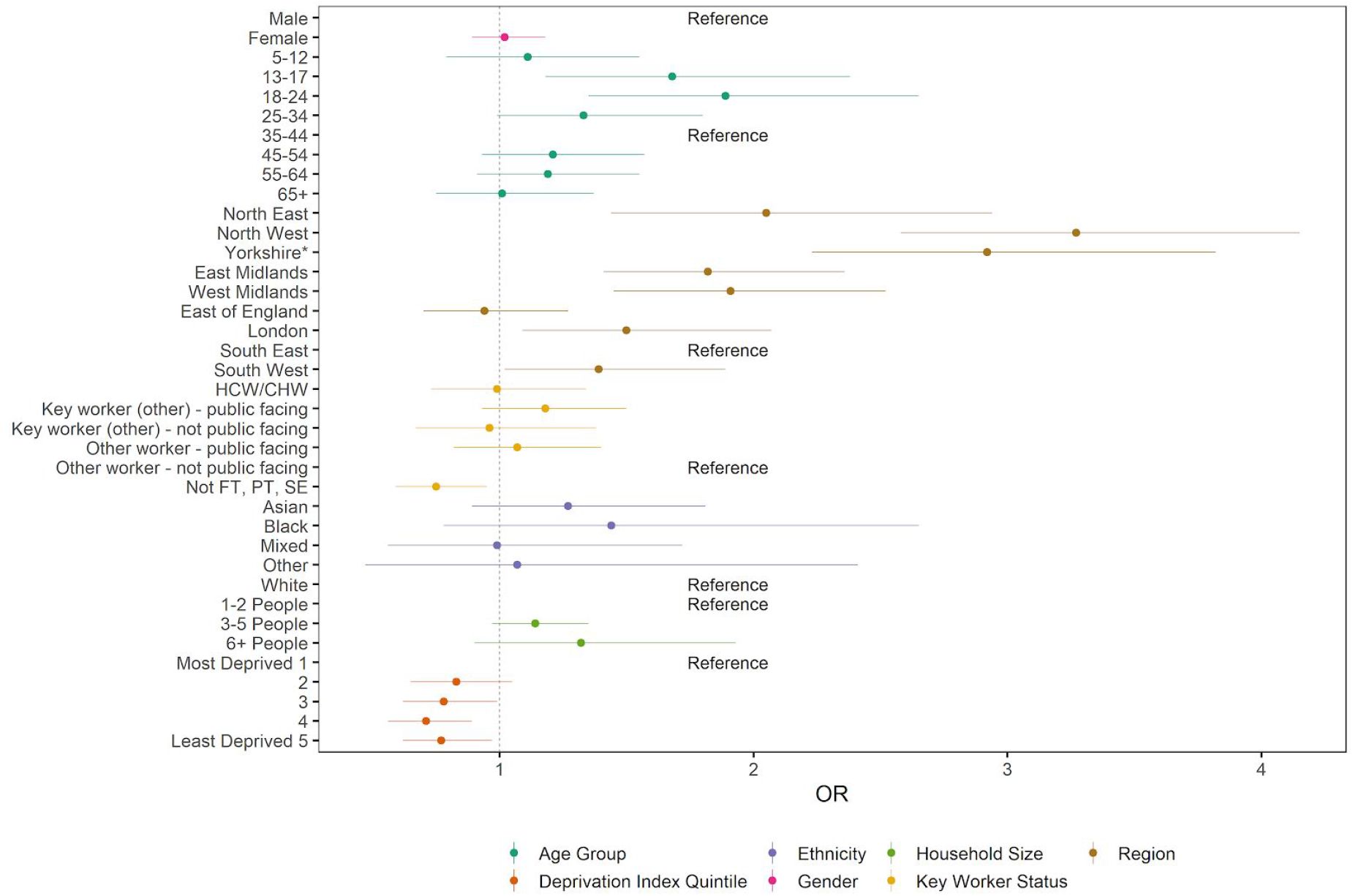
Estimated odds ratios and 95% confidence intervals for jointly adjusted logistic regression model of swab-positivity for round 6. The Deprivation Index is based on the Index of Multiple Deprivation (2019) at lower super output area. Here we group scores into quintiles, where 1 = most deprived and 5 = least deprived.

We observed decreased odds of swab positivity in individuals who do not work full-time, part-time, or are not self-employed at 0.64 (0.48, 0.85) compared to other workers with non-public facing rolls. We also observed decreased odds in individuals living in less deprived areas compared to those living in the most deprived areas (Table 5). When comparing odds ratios between the unadjusted, adjusted for age and gender, and jointly adjusted models, the odds ratios for most covariates went towards one as more variables were included in the models. However, odds ratios for age group and deprivation quintile went away from one as more covariates were included.

**Table 5.**
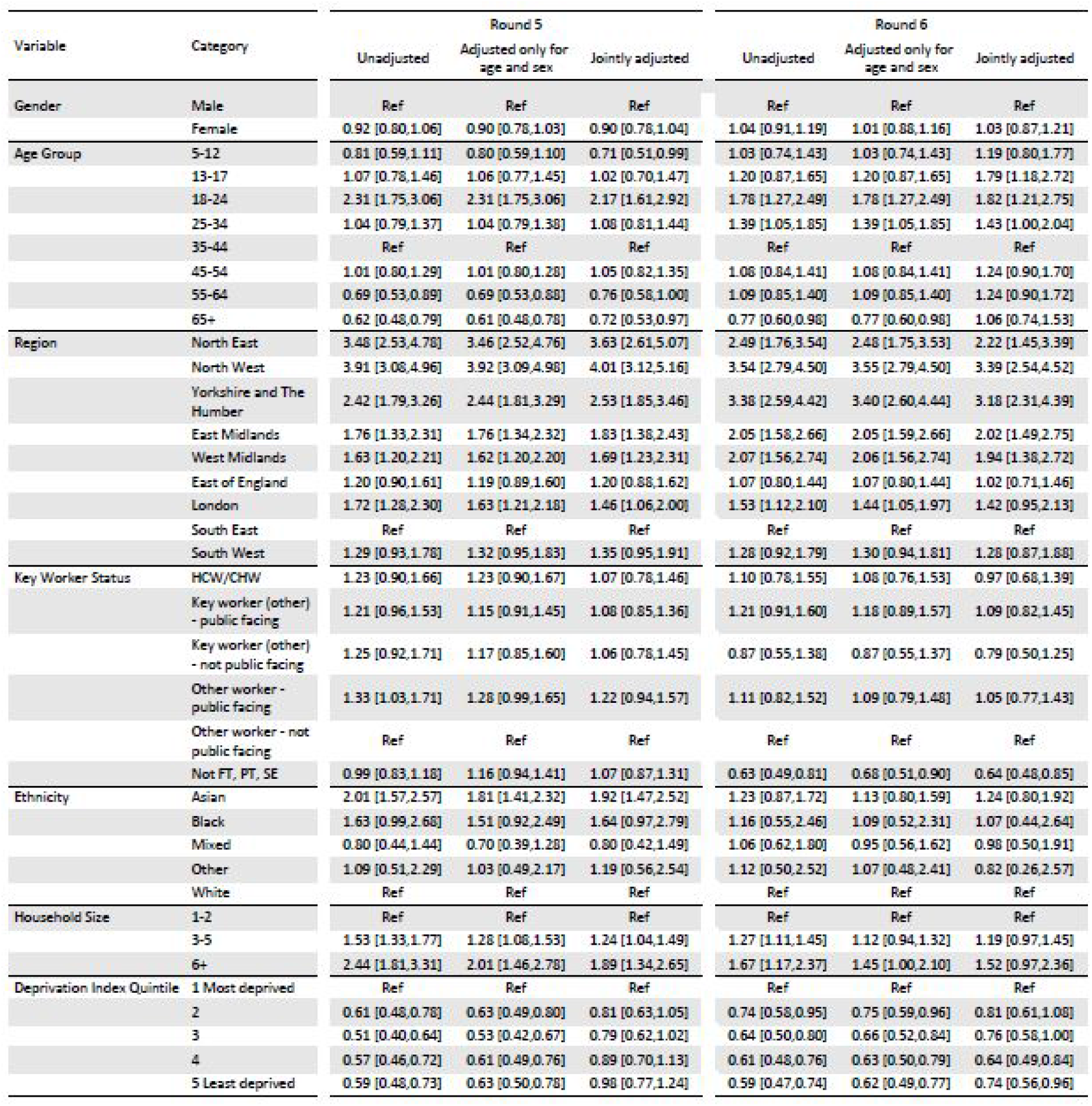
Estimated odds ratios for logistic regression models of swab-positivity for REACT-1 rounds 5 and 6.

Patterns of swab-positivity by age for rounds 5 and 6 were different across regions (Figure 7). In the highest prevalence regions of Yorkshire and The Humber and North West, there has been an apparent slowing or decline in 18 to 24 years olds but with evidence of more rapid growth in older people and school-aged children. Conversely, in the South West there has been a rapid increase in 18 to 24 year olds but to date slower growth in older age groups.

**Figure 7.**
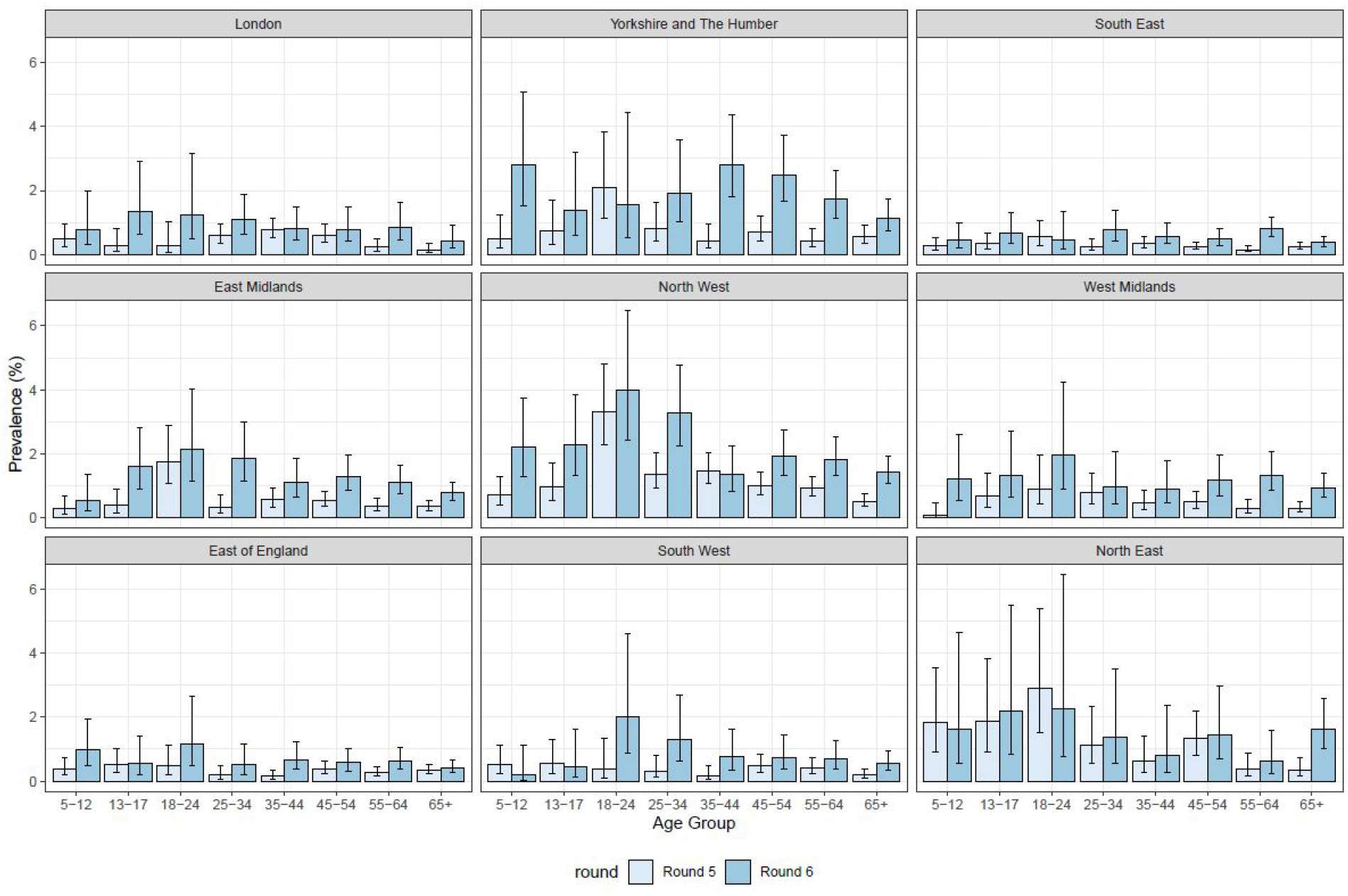
Weighted prevalence and 95% confidence intervals of swab-positivity by age and by region for rounds 5 and 6.

Figure 8 shows the jittered locations of the most clustered individuals testing positive in rounds 5 and 6 to date. In the North, there is strong clustering in Lancashire, Manchester, Liverpool, and West Yorkshire. Similar patterns can also be seen in Figure 9 which shows unweighted prevalence by lower-tier local authority.

**Figure 8.**
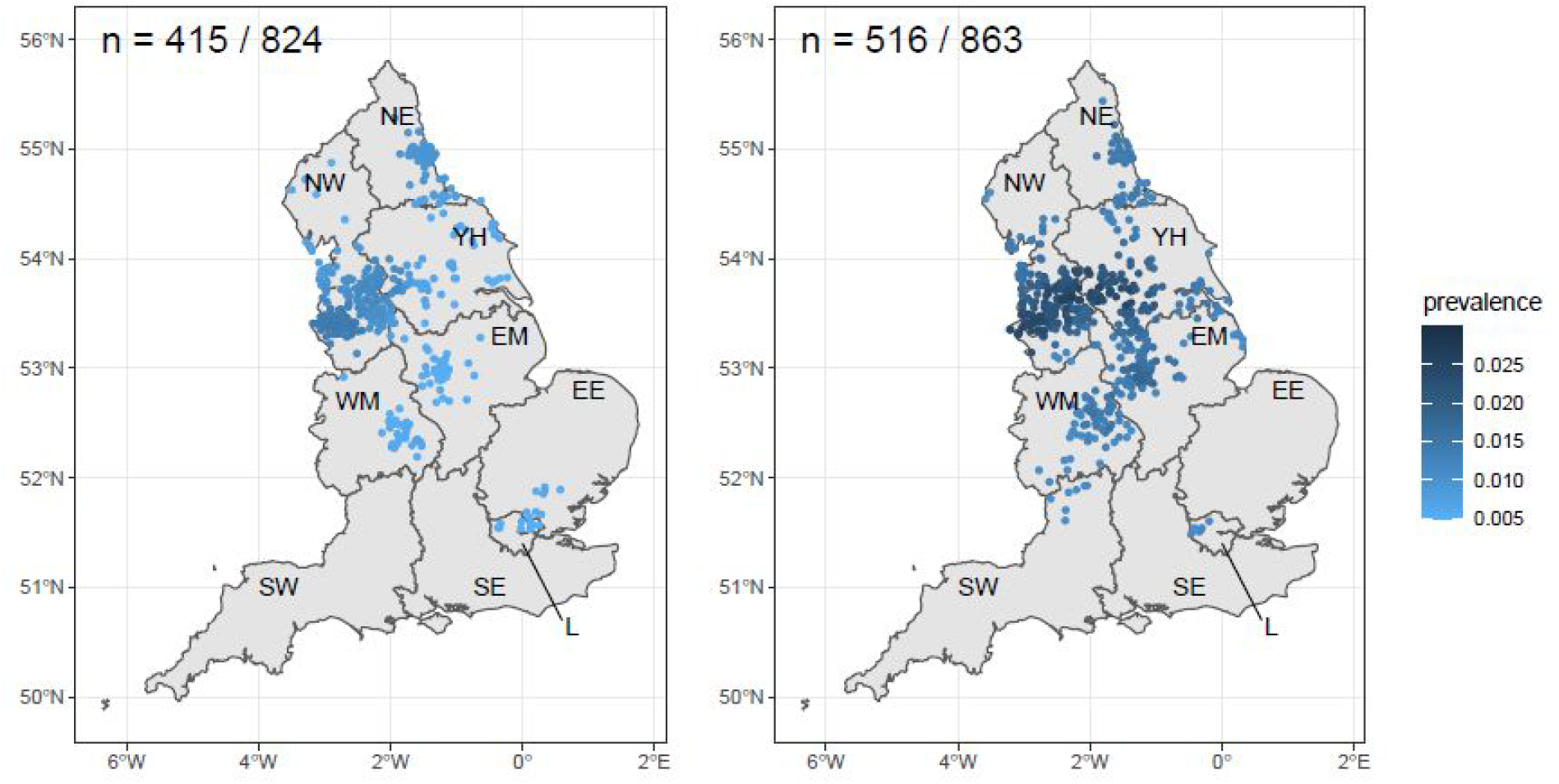
Jittered home locations of 415 (round 5, left) and 516 (round 6, right) swab-positive participants with high local prevalence. Prevalence calculated from nearest neighbours (the median number of neighbours within 30 km in the study). All points marked have estimated prevalence higher than the upper 95% confidence bound of national average. Regions: NE = North East, NW = North West, YH = Yorkshire and The Humber, EM = East Midlands, WM = West Midlands, EE = East of England, L = London, SE = South East, SW = South West. Because we sample from the neighbour distribution, independent runs of this algorithm give slightly different numbers of clustered points.

**Figure 9.**
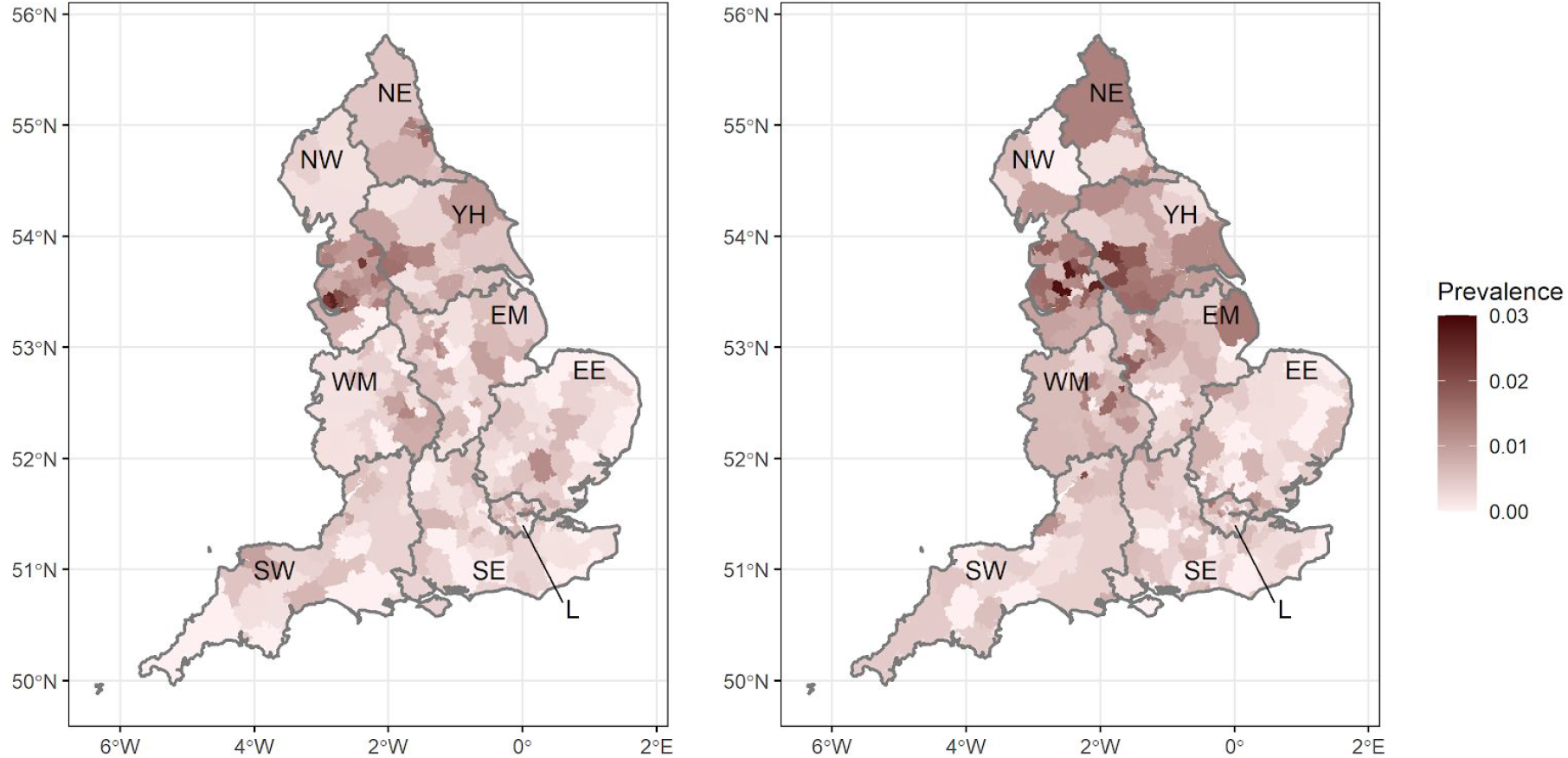
Prevalence of swb-positivity by lower-tier local authority for England for round 5 (left) and round 6 (right). Regions: NE = North East, NW = North West, YH = Yorkshire and The Humber, EM = East Midlands, WM = West Midlands, EE = East of England, L = London, SE = South East, SW = South West.

## Discussion

We first described a resurgence in SARS-CoV-2 infections in England in August-September 2020 [1] at the beginning of the second wave. Here we report an acceleration in infections in England during October 2020. The estimated reproduction number R has risen to 1.6 during October from 1.2 in the prior round of the study and infections have spread from the lower-risk 18 to 24 year old age group to older age groups including those most at risk, aged 65 years or older. We previously reported that prevalence was highest in northern parts of England [2]. While this is still the case, there is suggestion that the epidemic may be turning down in the North East, although there are still marked increases in prevalence amongst the most vulnerable population at ages 65 years and over. The epidemic is now increasing most rapidly in the Midlands and South. Patterns of growth rate and the age distribution of cases in the South now are similar to those observed in northern regions during the prior two rounds of this study.

A suite of intervention measures are currently in place across England, designed to reduce transmission while allowing continued social and economic activity. These measures include: the provision of free testing, mandated isolation and household quarantine, contact tracing and contact quarantining, identification and contact tracing of locations at which multiple infections have occurred, limits on the size of social gathering, and the mandated use of mask wearing in retail outlets and on public transport. Also, additional restrictions are in place in some geographical areas according to a tiering system, with hospitality businesses greatly restricted in the highest tier. However, our results suggest strongly that one or more of: the policies themselves, the timing of tier advancement, or levels of compliance, have not been sufficient to date to achieve control of the second wave of COVID-19 in England.

Based on our prevalence estimate at national level of 1.28%, we estimate that 960,000, (860,000, 1,050,000) individuals are harbouring SARS-CoV-2 virus in England on any one day. This assumes a 75% sensitivity for virus (if present) from a nose and throat swab [5,6]. With the further assumption that on average the virus can be detected for 10 days after initial infection, this corresponds to 96,000, (86,000, 105,000) new infections per day. Our data across all previous rounds of our study indicate that at least half of people with detectable virus will not report symptoms on the day of testing or week prior [2], therefore studies that rely only on symptomatic reporting of cases will under-estimate population incidence.

Our study has limitations. We assume that across each round the individuals taking part have similar characteristics including propensity to test positive or negative given the levels of circulating virus at the time. It is possible that symptomatic individuals may choose instead to be tested through the national “Test and Trace” system or alternatively may be more likely to participate in our study in order to obtain a test result. While both sources of bias are possible, we have no reason to believe they are operating to any large extent, and in any case would be unlikely to explain trends in our data within any one round. We also weight the data to be representative of England as a whole, which should correct for under- or over-representation of any particular group. Also, we rely on self-administered nose and throat swabs which may miss virus (if present) in 20% to 30% of people [6]. In addition, any changes in laboratory procedures may affect positivity rates. However, we have been using the same methods across all rounds of the study, and same laboratory and protocols (with strict quality control), which should minimise any such errors.

The co-occurrence of high prevalence and rapid growth means that the second wave of the epidemic in England has now reached a critical stage. While it is possible that some of the current control measures may be too recent to have fed through to the data reported here, the high prevalence already reached and the rapid acceleration mean that inevitably there will be large numbers of hospitalisations and deaths resulting from the second wave. Whether via regional or national measures, it is now time-critical to control the virus and turn R below one if yet more hospital admissions and deaths from COVID-19 are to be avoided.

## Data Availability

The datasets generated or analysed, or both, during this study are not publicly available because of governance restrictions.

## Acknowledgements

SR, CAD acknowledge support: MRC Centre for Global Infectious Disease Analysis, National Institute for Health Research (NIHR) Health Protection Research Unit (HPRU), Wellcome Trust (200861/Z/16/Z, 200187/Z/15/Z), and Centres for Disease Control and Prevention (US, U01CK0005-01-02). GC is supported by an NIHR Professorship. PE is Director of the MRC Centre for Environment and Health (MR/L01341X/1, MR/S019669/1). PE acknowledges support from Health Data Research UK (HDR UK), the NIHR Imperial Biomedical Research Centre and the NIHR HPRUs in Environmental Exposures and Health and Chemical and Radiation Threats and Hazards, the British Heart Foundation Centre for Research Excellence at Imperial College London (RE/18/4/34215) and the UK Dementia Research Institute at Imperial (MC_PC_17114). We thank The Huo Family Foundation for their support of our work on COVID-19.

We thank key collaborators on this work -- Ipsos MORI: Kelly Beaver, Sam Clemens, Gary Welch, Nicholas Gilby, Andrew Cleary and Kelly Ward; Institute of Global Health Innovation at Imperial College: Gianluca Fontana, Dr Hutan Ashrafian, Sutha Satkunarajah and Lenny Naar; MRC Centre for Environment and Health, Imperial College London: Daniela Fecht; North West London Pathology and Public Health England for help in calibration of the laboratory analyses; NHS Digital for access to the NHS register; and the Department of Health and Social Care for logistic support. SR acknowledges helpful discussion with members of the UK Government Office for Science (GO-Science) Scientific Pandemic Influenza – Modelling (SPI-M) committee.

## Tables and Figures

Supporting data for Tables and Figures are [here](https://docs.google.com/spreadsheets/d/1WNnVsEP3NLipDozliXJqtwhbgCHyfk_OE13F0-SPdZE/edit).

